# Quantitative Analysis of Ablation Technique Predicts Arrhythmia Recurrence Following Atrial Fibrillation Ablation

**DOI:** 10.1101/19013318

**Authors:** Lior Jankelson, Matthew Dai, Scott Bernstein, David Park, Douglas Holmes, Anthony Aizer, Larry Chinitz, Chirag Barbhaiya

**Affiliations:** Leon H. Charney Division of Cardiology, NYU Langone Medical Center, New York University School of Medicine, New York, USA

**Keywords:** Ablation, Stability, Excursion, Impedance

## Abstract

**Background:** Optimal ablation technique, including catheter-tissue contact during atrial fibrillation (AF) radiofrequency (RF) ablation is associated with improved procedural outcomes. We used a custom developed software to analyze high frequency catheter position data to study the interaction between catheter excursion during lesion placement, lesion-set sequentiality and arrhythmia recurrence.

**Methods:** 100 consecutive patients undergoing first time RF ablation for paroxysmal AF were analyzed. Spatial positioning of the ablation catheter sampled at 60 Hz during RF application was extracted from the CARTO3 system (Biosense Webster Inc., USA) and analyzed using custom developed MATLAB software to determine precise catheter spatial 3D excursion during RF ablation. The primary end point was freedom from atrial arrhythmia lasting longer than 30 seconds after a single ablation procedure.

**Results:** At one year, 86% of patients were free from recurrent arrhythmia. There was no significant difference in clinical, echocardiographic or ablation characteristics between patients with and without recurrent arrhythmia. Analyzing 15,356,998 position data-points revealed that lesion-set sequentiality and mean lesion catheter excursion were predictors of arrhythmia recurrence. Analyzing arrhythmia recurrence by mean single-lesion catheter excursion (excursion > 2.81mm) and by sequentiality (using 46% of lesions with inter-lesion distance >6mm as cutoff) revealed significantly increased arrhythmia recurrence in the higher excursion group (23% vs. 6%, p=0.03) and in the less sequential group (24% vs. 4%, p=0.02).

**Conclusion:** Ablation lesion sequentiality measured by catheter inter-lesion distance and catheter stability measured by catheter excursion during lesion placement are potentially modifiable factors affecting arrhythmia recurrence after RF ablation for AF.

## Background

Pulmonary vein isolation (PVI) is foundational to catheter ablation procedures for prevention of paroxysmal atrial fibrillation (AF)^1^. Optimal catheter-tissue contact-force (CF) during RF ablation is associated with reduction in arrhythmia recurrence following AF ablation, ^2-4^ which remains common despite utilization of force-sensing ablation catheters.^5^ While CF instability, often indirectly defined by average CF or standard deviation of CF is associated with poor lesion quality,^6^ The extent to which *spatial instability* - measured *directly* during single lesion RF application - contributes to ineffective lesion formation and arrhythmia recurrence is unknown. Similarly, it is unclear if differences in the sequentiality of lesion placement technique is impacting procedure outcomes. In the present study we retrospectively applied a custom developed computational tool to quantitatively analyze ablation catheter spatial position during RF ablation of paroxysmal AF, with the following aims: 1) evaluate the interaction between ablation catheter spatial position, both in terms of lesion-set sequentiality as well as stability during individual lesion placement, and the clinical outcomes of RF ablation in patients with paroxysmal atrial fibrillation, and 2) assess the interaction between spatial catheter stability and ablation parameters.

## Methods

### Patients and procedure

We retrospectively analyzed 100 consecutive patients undergoing first time RF ablation for paroxysmal atrial fibrillation in our center, included in our prospective AF ablation registry. All procedures were performed under general anesthesia with mechanical ventilation and using respiratory gating to adjust positional data for respiratory motion. Patients were ablated in sinus rhythm with atrial pacing at 600 ms cycle length to enhance catheter stability. Surface and intracardiac electrocardiograms were recorded and stored (EP Workmate, St. Jude Medical, Inc., USA). A detailed left atrial electroanatomical map in sinus rhythm was created using a circular mapping catheter or a five-spline mapping catheter (Lasso or PentaRay, Biosense Webster, Inc., USA) and 3-D electroanatomic mapping system (CARTO3, Biosense Webster, Inc. USA). Wide antral PVI was then performed using a 3.5mm tip force-sensing RF ablation catheter (ThermoCool SmartTouch, Biosense Webster, Inc., USA), set to 35 Watts for a force-time integral value of 400 g*s on the anterior wall and 200 g*s on the posterior wall. Lesions were placed using point-by-point technique with no dragging allowed. No visual gaps in the PVI were allowed in the end of the case. ViziTag lesion settings were set to 2mm/5sec motion and 5 gr of minimal force. Esophageal temperature probe was used in all patients for monitoring (Truer Medical, USA). Electrical isolation of the pulmonary veins, both entrance and exit block, was confirmed using pacing and Adenosine administration.

### Ablation Data analysis

Data was extracted from the electroanatomical mapping system and analyzed using custom MATLAB software (Mathworks, USA). Catheter 3D position and impedance sampled at 60Hz were registered for each ablation lesion. Patient specific spatial catheter stability, represented by catheter excursion was determined as follows: for each ablation lesion, continuous 3D catheter position was extracted and the displacement of the catheter tip relative to the center of each lesion was determined (Figure 1). Catheter excursion was defined as following: maximal excursion was defined as the farthest distance from the center of a single ablation lesion, which the catheter traveled during a single lesion. Mean excursion was defined as the average distance the catheter from the center of the lesion during a single ablation lesion. Catheter stability was then calculated for each case as the average excursion (maximal and mean, separately) during all RF lesions applied during a single procedure in a single patient. To study the implication of lesion placement-order on ablation outcomes, we defined the variable “lesion-sequentiality” (Figure 1D). In each patient, we calculated the percent of temporally sequential lesion pairs (i.e. 2 lesions place immediately one after another), which were spatially non-sequential, i.e. placed greater than 6 mm apart. This variable represents the order of lesion placement: in cases where the operator tended to move the catheter more frequently between different atrial locations, lesion sequentiality was lower (more temporally sequential lesion pairs were placed >6mm apart). Regardless of lesion sequentility, all lesion sets in all patients were complete, with lesions overlapping and no gaps permitted along the PVI lines.

**Figure 1.** (A-D): A, catheter motion during a single RF lesion sampled at 60Hz is represented by the dotted fine pink line, each dot representing a discrete sampled position. Outer blue line represents maximal catheter excursion (the farthest point from the center of the lesion which the catheter reached during single lesion placement) and the dashed green line represents mean catheter excursion (average distance from the center of the lesion during lesion placement). The PVI lesion maps of unstable (B) and stable (C) ablation cases are presented. In these panels, mean catheter excursion is represented by sphere size, where a smaller sphere represents a more stable lesion. Note the larger spheres evident in the unstable relative to the stable map. (D) illustration of lesion sequentiality. The posterior left atrium is presented, with 2 lines consisting of 4 ablation lesions, numbered by their order of placement. In the left line, lesions were placed sequentially: the distance between any 2 sequential lesions (i.e. 1 to 2, 2 to 3, 3 to 4) is always < 6mm. In the right ablation line, lesions were placed non-sequentially: lesion 2 was placed > 6mm away from lesion 1, and lesion 3 was placed > 6mm away from lesion 2. Note that in both cases there are no gaps in the lesion-sets.

### Clinical Endpoint

All patients underwent two-week event-monitoring at 3, 6 and 12 months post procedure. The primary endpoint of the study was freedom from AF lasting more than 30 seconds, following a 3-month blanking period^1^.

### Statistical analysis

All statistical analysis was performed using SPSS Statistics 23 (SPSS Inc., USA) and graphs were constructed using Prism 7 (GraphPad Software, Inc., USA). Continuous variables were expressed as either mean ± standard deviation for tables or mean ± standard error of the mean for figures. Categorical variables were expressed as percentages. Normality of samples with continuous variables was assessed using Shapiro-Wilk test. Two sample hypotheses testing for continuous variables was performed using either Student’s *t*-test if samples had normal distributions or Mann-Whitney *U* test if samples did not have normal distributions. Hypothesis testing with multiple samples was performed using non-parametric Kruskal-Wallis test with subsequent pairwise comparison using Mann-Whitney *U* test and Dunn-Bonferroni correction to significance level α = 0.05. For non-composite data point comparisons performed, we used an additional mixed-design analysis of variance (split-plot ANOVA) to account for both random and fixed effects introduced by clusterization. For categorical variables, hypothesis testing was performed using Fisher’s exact test. Logistic regression was performed to identify predictors of arrhythmia recurrence. This approach was also performed to identify predictors of catheter instability. For Kaplan-Meier survival analysis, the median maximal excursion (2.81mm) was chosen as the cut-off value to divide the dataset into two equal groups. Significance analysis between groups was performed using log-rank test.

All patients provided written informed consent for their procedures, and all data collection and analysis were performed in accordance with the NYU Langone Medical Center Institutional Review Board.

No extramural funding was used to support this work. The authors are solely responsible for the design and conduct of this study, all study analyses, the drafting and editing of the paper and its final contents

## Results

### Patient characteristics and procedural outcomes

Of 100 consecutive cases included, fourteen (14%) had continuous implantable monitoring with either a PPM (n=7), or implantable loop recorder (n=7). The remaining patients received an average of 2.1±1.0 two-week monitors during the follow-up period. Patient characteristics are presented in table 1.

At 1-year follow-up, overall Kaplan-Meier estimated arrhythmia free survival was 86%. There were no differences in baseline characteristics, including age, gender, EF and LA size, between patients with and without arrhythmia recurrence.

A total of 14,703 ablation lesions with an average of 1044±525 position data-points per lesion were studied, for a total of 15,356,998 position data-points. Mean catheter excursion and maximal catheter excursion were significantly greater in the recurrence group versus the arrhythmia-free group (1.08±0.13mm vs. 0.98±0.17mm p=0.01 and 3.07±0.38mm vs. 2.80±0.40mm respectively, p=0.03). Percent lesions with inter-lesion distance > 6mm was significantly higher in the arrhythmia recurrence group (53±9% vs. 46±8, P<0.001), Figure 2. There were no differences between groups in all other ablation parameters (Table 2), including number of PVs, RF time, average power delivered, ablation catheter average CF or standard deviation of CF, lesion duration, FTI or impedance decline.

**Figure 2.** (A) mean excursion by arrhythmia recurrence and (B) mean interlesion distance by arrhythmia recurrance. \**p* < 0.01 by Mann-Whitney U test. Dashed line represents the mean value in the cohort.

### Predictors of arrhythmia recurrence

Significant predictors of arrhythmia recurrence by logistic regression (Table 3) were greater percent lesions with >6mm inter-lesion distance, increased catheter excursion, (for maximal catheter excursion OR 5.15 per 1mm maximal excursion increase, 95% CI 1.21-21.9, p=0.04) and lower ejection fraction.

### Arrhythmia recurrence by spatial catheter excursion

Maximal catheter excursion (meadian, 2.81mm) and lesion sequentiality (median percent lesion pairs with > 6mm inter-lesion distance) were used separately to evaluate arrhythmia recurrence (figure 3). Kaplan-Meier estimated arrhythmia free survival was significantly greater in patients with lesser than median maximal catheter excursion, compared to patients with greater than median maximal catheter excursion (94% vs. 77% respectively, p=0.02). Arrhythmia free survival was significantly higher in patients with greater lesion sequentiality, evident by smaller percent of lesion pairs > 6mm apart (96% vs. 76%, p=0.01).

**Figure 3.** Kaplan Meier arrhythmia-free survival curve, separated by (A) median maximal catheter excursion (2.81mm) and by (B) percent of lesion pairs with > 6mm interlesion distance.

**Predictors of ablation catheter spatial stability**

Comparing clinical and ablation parameters categorically using median maximal excursion (2.81mm) as a cutoff for defining catheter stability, increased body surface area (BSA), increased body weight and the presence of CHF were associated with greater catheter excursion (Supplemental Table 1). Logistic regression demonstrated that decreased body weight and LA size were significant predictors of catheter stability, as well as ablation by specific operators (Supplemental Table 2). Ablation parameters associated with better catheter spatial stability were decreased lesion number, longer lesion duration and lower percent lesions with >6mm inter-lesion distance. No differences in CF, standard deviation of CF, FTI, Impedance decline or total RF time were observed between groups. No association was found between CF and catheter stability when assessed by quartiles of CF.

### Catheter spatial stability and impedance

Spatial stability, FTI, CF and standard deviation of CF all demonstrated poor correlation with impedance decline (r = 0.15, 0.14,0.07 and 0.02 respectively, p < 0.001 for all pairs). Figure 4 illustrates an example of closely-spaced pair of lesions, where large differences in spatial stability and impedance decline are observed, without significant difference in CF.

**Figure 4.** (A-D): typical example from a patient undergoing AF ablation. In A, the right PV lesion set is shown. B, 2 lesions with inter-lesion distance of 5.21mm are magnified. Note the differences in spatial instability, represented by sphere diameter. C, Larger impedance decline is noticed in lesion A relative to B, despite lesion B being more stable. D, the nearly 2-fold impedance decline in lesion A vs. B is accompanied by high and overlapping CF in both lesions.

## Discussion

In this study of 100 consecutive patients undergoing RF ablation for paroxysmal AF, we found that:

1. Predictors of arrhythmia recurrence following paroxysmal AF ablation were:
  a. Ablation catheter single lesion spatial stability during lesion placement, measured using high frequency catheter excursion data
  b. Lesion sequentiality, measured by percent of temporally sequential lesion pairs placed > 6mm apart, and
  c. lower ejection fraction
2. Impedance decline during RF lesion delivery was poorly associated with ablation parameters, including catheter stability and contact-force.

### Ablation catheter spatial stability and lesion-set sequentiality predict arrhythmia recurrence

In the present work, we studied ablation catheter quantitative spatial stability, which we defined and evaluated by analyzing over 15 million catheter position data-points stored by the electroanatomical mapping system. The ability to precisely analyze positioning data is possible due to high spatial resolution of the mapping system, less than 1mm^7^ over the entire applied field. We used custom MATLAB code applied over CARTO system output to rapidly extract, analyze and graphically present ablation catheter positional data. We show that using a more sequential technique in placing the ablation lesions is associated with reduced arrhythmia recurrence. We also demonstrate that relatively small differences in catheter excursion during lesion placement are significant in predicting arrhythmia recurrence. We found that the clinical variables associated with diminished catheter stability are increased body weight and the presence of CHF. We did not find significant differences in potential modifiers of ablation outcome such as the number of lesions, average force or FTI, between patients with and without recurrence. It is possible that increased cardiac motion associated with increased cardiac size, as seen in both patients with higher body weight as well as CHF, translates to more significant catheter motion. The procedural markers characterizing cases with diminished catheter stability were less sequential lesion-set, increased number of lesions and shorter lesion duration. To date, in both pre-clinical ^6, 8^ and clinical studies, catheter stability was assessed indirectly using CF variability (CF standard deviation) which was found to be associated with suboptimal lesion formation and clinical outcomes^9, 10 11^. However, irrespective of CF and its variability, cardiac and respiratory motion may result in *spatial* displacement of the ablation electrode relative to underlying tissue. This spatial displacement of the ablation electrode, which may not be reflected in CF variability, may impact lesion quality and increase the likelihood of arrhythmia recurrence. Concordant with our findings, in a post-hoc analysis of automated lesion annotation in the SMART-AF study, gaps between lesion markers created using strict stability criteria predicted arrhythmia recurrence^12^. While we identified increased BSA (driven by body weight) as predictor of catheter spatial instability, it did not predict arrhythmia recurrence. In the analysis of ablation parameters by catheter stability (rather than by arrhythmia recurrence), cases with lower stability were associated with less sequential lesion-set, greater lesion number and shorter lesion duration. This finding may imply that operators were aware and responded to catheter instability. Indeed, improving spatial catheter stability may be a mechanism by which interventions such as high frequency jet ventilation and the use of a deflectable sheath improve clinical outcomes^13-15^. Additional proposed measures that may improve spatial catheter stability during RF application are rapid pacing and the use of periodic apnea^16^. The rhythm during AF ablation is another modifier of catheter stability recently shown to modulate catheter contact force, and may be a potential strategy for increasing catheter stability^17^. The optimal means by which to evaluate and modify intra-procedural spatial catheter instability require further evaluation.

### Impedance change during RF ablation

Decrease in the unipolar impedance of the ablation electrode circuit during RF application is widely used as surrogate marker for effective lesion creation. This, is based on both pre-clinical and clinical studies demonstrating a positive correlation between the magnitude of impedance decline and CF, lesion size, and ablation outcomes^18, 19 20^. However, the strength of correlation between CF and impedance decline varies widely between clinical studies ^21, 22^, and over the narrow range of CF observed in the present analysis, we found the correlation to be weak. In this perspective, previously unacknowledged variability in spatial stability may help explain the large discrepancies in correlations of impedance decline and CF reported in previous studies, where wide range of impedance decline was recorded over different ranges of CF^21, 23^.

Taken together, these data suggest that the factors that contribute to impedance decline during RF application remain incompletely understood.

### Limitations

This study was a retrospective analysis of a prospective registry enrolling all patients undergoing AF ablation at our institution; this however, is partially mitigated by the all-comer, consecutive nature of the study, offsetting the potential for selection bias. We were not able to assess first-pass PV isolation nor the cause of arrhythmia recurrence in the 14 patients with post ablation arrhythmia, and it is possible that the mechanism of recurrence was non-PV related. However, inadequate lesion quality may create the relevant substrate for both PV reconnection as well as for non-PV related tachyarrhythmia, including AF and AT. Further assessment of the mechanism of recurrence may be addressed in a future study. Although significantly different and predictive, values of catheter stability and sequentiality demonstrated overlap between the groups with and without recurrence. Hence, a prospective study on a larger population would be required to validate our findings. Lastly, 14 patients were monitored with ILRs, while 86 patients were monitored with conventional, guideline-directed Holters. Although arrhythmia recurrence and ablation parameters did not vary between patients with and without an ILR, under-detection of arrhythmic events is possible.

## Conclusion

Ablation catheter spatial instability and decreased lesion sequentiality are predictors of arrhythmia recurrence in patients undergoing RF ablation for paroxysmal AF, separately resulting in nearly 3-fold risk of arrhythmia recurrence. Additional studies are required to further define the importance of quantitative real-time monitoring of catheter stability and lesion placement technique, and the utility of interventions aimed at improving them.

## Data Availability

Data is available

## Acknowledgments

The authors would like to thank Mr. Morris Ziv-Ari (Biosense Webster Inc., USA) for his assistance with MATLAB scripting.

